# Durability of the insecticidal activity of next-generation insecticide treated nets distributed for malaria control in Mozambique: findings from the New Nets Project (2020–2022)

**DOI:** 10.1101/2025.09.24.25336605

**Authors:** Josias Fagbohoun, Ana Paula Abílio, Boris N’Dombidje, Damien Todjinou, Marie Baes, Olivier Pigeon, Germain Gil Padonou, Christen Fornadel, Baltazar Candrinho, Hannah Koenker, Molly Robertson, Joseph Wagman, Corine Ngufor

**Author notes:** Current Affiliation: The Global Fund, Geneva, Switzerland (Molly Robertson). Corresponding author CN.

## Abstract

**Background:** As next-generation insecticide-treated nets (ITNs) are increasingly deployed, it is essential to monitor their durability over time under operational conditions to support evidence-based procurement and replacement strategies by national malaria programs. This study evaluated the insecticidal durability of next-generation ITNs distributed in Mozambique through the New Nets Project (2020–2022), with a focus on bioefficacy and chemical content over 24 months of household use.

**Methods:** An ITN insecticidal durability study was conducted with nets from four districts of Mozambique to evaluate four ITN product types: pyrethroid-only (MAGNet^®^, DuraNet^®^, Olyset^®^ Net), pyrethroid-PBO (Olyset^®^ Plus), pyrethroid-pyriproxyfen (Royal Guard^®^), and pyrethroid-chlorfenapyr (Interceptor^®^ G2). ITNs were withdrawn from households at 6, 12, and 24 months post-distribution. Bioefficacy was assessed via WHO cone and tunnel bioassays using insecticide-susceptible and pyrethroid-resistant laboratory strains of *Anopheles gambiae* s.l., and through experimental hut trials in Cove, Benin against wild, pyrethroid-resistant *An. gambiae* s.l. populations. Chemical analysis was conducted to quantify active ingredient content over time.

**Results:** The pyrethroid components of all ITN types retained high efficacy, with 100% of nets meeting WHO bioefficacy criteria at 6, 12, and 24 months in bioassays using the susceptible *An. gambiae* KISUMU strain. However, the proportion of Olyset^®^ Plus nets meeting WHO thresholds in bioassays assessing the bioefficacy of the PBO component against the pyrethroid-resistant *An. gambiae* s.l. AKRON strain, declined from 73% at 6 months to 40% at 24 months. Royal Guard^®^ showed variable performance for pyriproxyfen bioefficacy, with pass rates increasing from 43% to 87% over the same period. Interceptor^®^ G2 maintained high bioefficacy against the pyrethroid-resistant VKPER strain, with ≥92% of nets meeting WHO efficacy criteria across all time points. Experimental hut trials confirmed the superior field performance of Interceptor^®^ G2 compared to pyrethroid-only nets, though mortality declined from 55% to 39% and blood-feeding inhibition from 72% to 19% between 6 and 24 months. Chemical analysis showed substantial degradation of PBO and chlorfenapyr by 24 months, with residual levels at 8% and 32% of baseline, respectively.

**Conclusion:** Next-generation ITNs provide improved protection against pyrethroid-resistant malaria vectors, but the durability of their enhanced efficacy varies by active ingredient. Interceptor^®^ G2 demonstrated the most consistent performance over 24 months, reinforcing its utility for control of pyrethroid resistant malaria vectors. However, declining personal protection and insecticide content underscore the need for long-term monitoring. These findings support the implementation of extended, context-specific durability studies to guide ITN selection, replacement strategies, and policy decisions for sustained malaria control.

## Introduction

Insecticide-treated nets (ITNs) are the most widely used intervention for malaria prevention in endemic regions and have played a critical role in the substantial reductions in malaria incidence and mortality observed over the past two decades [1]. However, the emergence and spread of resistance to pyrethroids, the primary class of insecticides used on ITNs, has increasingly threatened their effectiveness, particularly in high-transmission areas [2-4]. To address this challenge, new generations of ITNs incorporating insecticide synergists or additional active ingredients have been developed to restore and enhance vector control in the face of resistance [5]. These include pyrethroid-PBO nets, which combine a pyrethroid insecticide with the synergist piperonyl butoxide (PBO) to inhibit key metabolic resistance mechanisms in mosquitoes [6, 7], and dual active ingredient nets that combine a pyrethroid with either pyriproxyfen [8, 9], an insect growth regulator that sterilizes mosquitoes, or chlorfenapyr, a pyrrole insecticide with a novel mode of action targeting mosquito mitochondria [10, 11]. These innovations aim to sustain the effectiveness of ITNs and prolong their impact on malaria transmission in regions where conventional pyrethroid-only nets are becoming less effective.

Pyrethroid-PBO nets were the first next-generation ITNs to demonstrate improved entomological and epidemiological efficacy over pyrethroid-only nets in multiple studies across sub-Saharan Africa, leading to their recommendation by the World Health Organization (WHO) in 2017 for deployment in areas of moderate pyrethroid resistance [12-14]. More recently, based on results from large cluster-randomized controlled trials conducted in Burkina Faso [15], Benin [16] and Tanzania [17], in 2023 the WHO issued a strong recommendation for the deployment of pyrethroid-chlorfenapyr nets and a conditional recommendation for pyrethroid-pyriproxyfen nets in areas of widespread pyrethroid resistance [5], following evidence of their improved protection against clinical malaria compared to standard pyrethroid-only ITNs. As the deployment of next-generation ITNs expands, there is an urgent need to monitor their survivorship, fabric integrity, insecticidal activity, and chemical content over time under operational conditions to support evidence-based procurement and replacement strategies by national malaria control programs [18]. This includes the development and adoption of standardized, high-throughput methods, as well as the identification of appropriate mosquito strains, for assessing the insecticidal durability of non-pyrethroid compounds such as chlorfenapyr and pyriproxyfen—areas that are not yet fully addressed by existing WHO guidelines for such studies.

The New Nets Project (NNP), launched in 2019, was designed to accelerate the introduction of next-generation ITNs while generating critical evidence on their effectiveness and durability under real-world conditions [19, 20]. As part of this initiative, a study was conducted with nets obtained from Mozambique between 2020 and 2022 to evaluate the bioefficacy and chemical retention of several next-generation and standard ITNs distributed to communities. Nets retrieved from households at 6, 12, and 24 months post-distribution were assessed for insecticidal activity through laboratory bioassays using both insecticide-susceptible and insecticide-resistant laboratory strains of *Anopheles gambiae* s.l., as well as in experimental huts at Covè, Benin, against wild, free-flying pyrethroid-resistant *An. gambiae* s.l. populations. In addition, chemical analysis of the nets was performed at baseline and at 12 and 24 months post-distribution to measure the retention of active ingredients over time.

## Materials and methods

### Study site and retrieval of study nets

The study sites in Changara, Gurue, Guro, and Mandimba districts of Mozambique (Figure 1) have been described in detail previously [19]. Cross-sectional sampling of ITNs distributed during the 2020 mass campaign was carried out over two years by the National Malaria Control Program and the National Health Institute with support from Tropical Health under the New Nets Project. Household sampling methods broadly followed the US President’s Malaria Initiative ITN durability monitoring protocol available at www.durabilitymonitoring.org.

**Figure 1.**
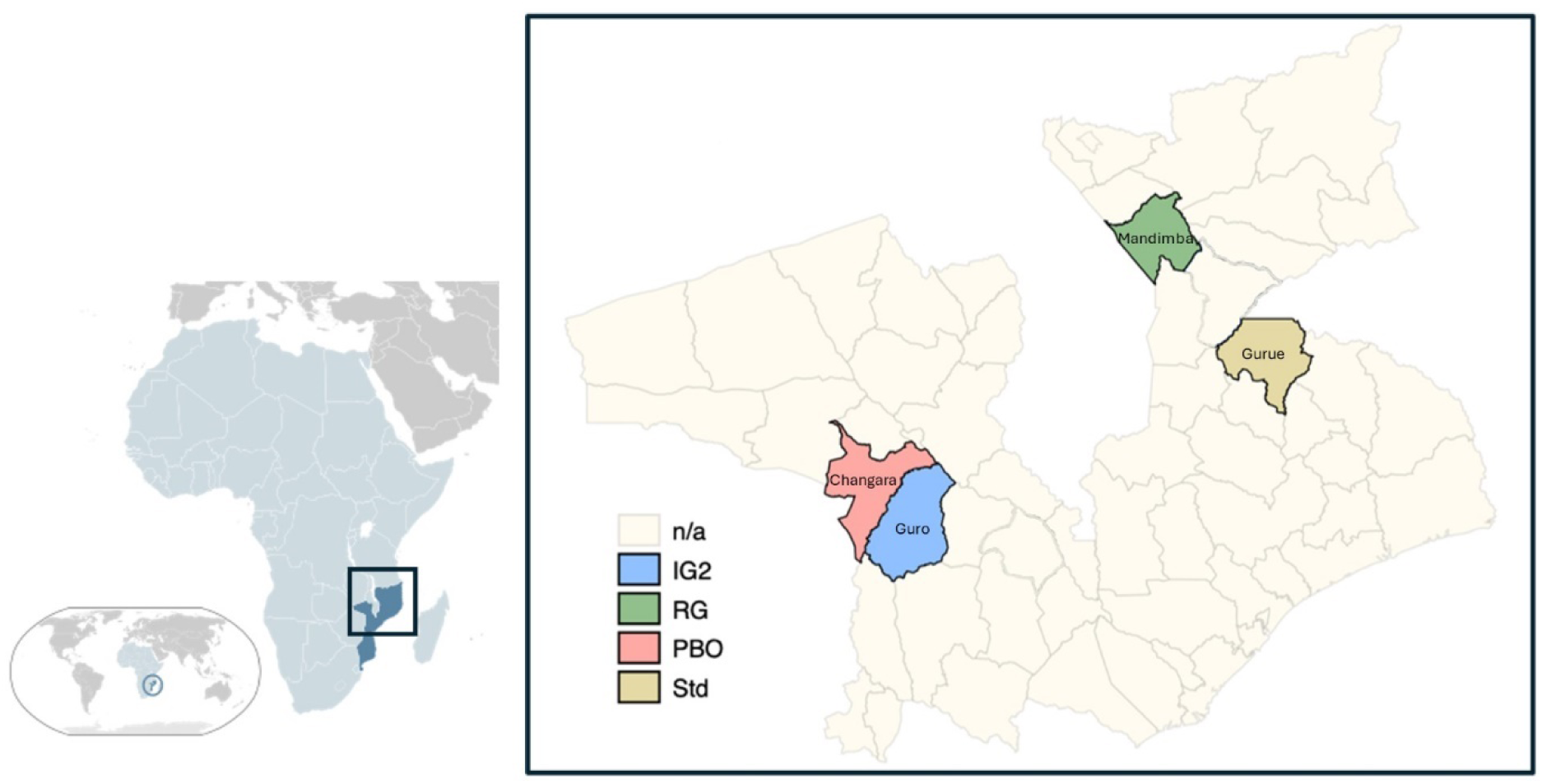
Study districts included in the durability evaluation of next-generation insecticide-treated nets (ITNs) distributed in Mozambique under the New Nets Project (2020–2022). *The map shows the geographic location of the study sites within Mozambique and the distribution of intervention arms across selected districts. Coloured shading indicates the ITN product evaluated in each district: Interceptor^®^ G2 (IG2, blue), Royal Guard^®^ (RG, green), Olyset^®^ Plus (PBO, red), and standard pyrethroid-only nets (Std, yellow)*.

At baseline, an ITN durability monitoring cohort in each district was established by selecting a representative sample of clusters (communities) based on probability proportionate to size and households using simple random sampling from household lists established on the day of the survey. At each timepoint, randomly selected campaign nets were retrieved from households neighboring study cohort households, and replaced with a new (unopened) ITN of the same type. ITN withdrawal at 6 months took place in March 2021, at 12-month in October 2021, and at 24 months in August 2022.

A description of the different types of ITNs withdrawn and assessed for bioefficacy and chemical content in this study is provided below:

a. Pyrethroid-only ITNs:
  - MAGNet^®^ (*V.K.A. Polymer Ltd*): Polyethylene net incorporating alpha-cypermethrin at 5.8 g/kg (±25%).
  - DuraNet^®^ (*Shobikaa Impex Pvt Ltd*): Polyethylene net incorporating alpha-cypermethrin at 5.8 g/kg (±25%).
  - Olyset^®^ Net (*Sumitomo Chemical Co., Ltd*): Polyethylene net incorporating permethrin at 20 g/kg (±25%).
b. Pyrethroid-PBO ITN:
  - Olyset^®^ Plus (*Sumitomo Chemical Co., Ltd*): Polyethylene net incorporating permethrin at 20 g/kg (±25%) and piperonyl butoxide (PBO) at 10 g/kg (±25%).
c. Pyrethroid-pyriproxyfen ITN:
  - Royal Guard^®^ (*Disease Control Technologies LLC*): Polyethylene net incorporating alpha-cypermethrin at 5.5 g/kg (±25%) and pyriproxyfen at 5.5 g/kg (±25%).
d. Pyrethroid-chlorfenapyr ITN:
  - Interceptor^®^ G2 (*BASF SE*): Polyester net coated with alpha-cypermethrin at 2.4 g/kg (±25%) and chlorfenapyr at 4.8 g/kg (±25%).

### ITN bioefficacy and chemical analysis

Table 1 presents the number of whole nets collected from the four study districts (Gurue, Changara, Mandimba, and Guro) for each ITN type and time point, along with the number of net pieces tested in cone or tunnel bioassays and those analysed for chemical content. Although some variation in the availability of pyrethroid-only ITNs was observed, particularly across brands, the sample sizes for next-generation ITNs (Olyset^®^ Plus, Royal Guard^®^, and Interceptor^®^ G2) remained consistent throughout. For all ITN types except Interceptor^®^ G2 at 6 and 24 months, net pieces measuring 30 × 30 cm were cut from whole nets at each time point in accordance with WHO guidelines, before being shipped to Benin. Two sets of pieces were prepared; one for bioefficacy testing and another for chemical analysis. All pieces were wrapped in aluminium foil and stored at ambient room temperature before and between bioassays. In contrast, for Interceptor^®^ G2 at 6 and 24 months, whole nets were shipped to Benin and first tested in experimental hut trials. Interceptor^®^ G2 net pieces for tunnel tests and chemical analysis were obtained after hut trial completion.

**Table 1:** Number of Interceptor^®^ G2, Royal Guard^®^, Olyset^®^ Plus and pyrethroid-only nets (Olyset^®^, MAGNet^®^ and DuraNet^®^) Interceptor^®^ ITNs tested for bioefficacy and chemical content.

| ITN type | Sampling site | Brand | No. of whole nets | No. of net pieces tested in bioassays | No. of net pieces analysed for chemical content |
| --- | --- | --- | --- | --- | --- |
| **6 months post-distribution** |  |  |  |  |  |
| Pyrethroid-only | Gurue | MAGNet® | 16 | 80 | 80 |
|  |  | DuraNet® | 5 | 25 | 25 |
|  |  | Olyset® Net | 9 | 45 | 45 |
| Pyrethroid-PBO | Changara | Olyset® Plus | 30 | 150 | 150 |
| Pyrethroid-pyriproxyfen | Mandimba | Royal Guard® | 30 | 150 | 150 |
| Pyrethroid-chlorfenapyr | Guro | Interceptor® G2 | 30 | 30 | 150 |
| **12 months post-distribution** |  |  |  |  |  |
| Pyrethroid-only | Gurue | MAGNet® | 22 | 110 | 110 |
|  |  | DuraNet® | 1 | 5 | 5 |
|  |  | Olyset® Net | 7 | 35 | 35 |
| Pyrethroid-PBO | Changara | Olyset® Plus | 30 | 150 | 150 |
| Pyrethroid-pyriproxyfen | Mandimba | Royal Guard® | 30 | 150 | 150 |
| Pyrethroid-chlorfenapyr | Guro | Interceptor® G2 | 30 | 30 | 150 |
| **24 months post-distribution** |  |  |  |  |  |
| Pyrethroid-only | Gurue | MAGNet® | 19 | 95 | 95 |
|  |  | DuraNet® | 4 | 20 | 20 |
|  |  | Olyset® Net | 7 | 35 | 35 |
| Pyrethroid-PBO | Changara | Olyset® Plus | 30 | 150 | 150 |
| Pyrethroid-pyriproxyfen | Mandimba | Royal Guard® | 31 | 155 | 155 |
| Pyrethroid-chlorfenapyr | Guro | Interceptor® G2 | 30 | 30 | 150 |

### Bioefficacy testing methods, mosquito strains, and efficacy criteria by ITN type

In line with current WHO guidelines [21], ITN bioefficacy at each time point was assessed by calculating the proportion of nets that met predefined efficacy thresholds for each active ingredient, based on results from cone bioassays or tunnel tests. Testing methods and mosquito strains were selected according to the insecticide class and its mode of action (see Table 2) in line with a similar New Nets Project study performed in Benin [18]. For pyrethroid-only nets (MAGNet^®^, Olyset^®^ Net, and DuraNet^®^), 3-minute cone bioassays were conducted using unfed 3- to 5-day-old *An. gambiae* (KISUMU strain) mosquitoes to measure knockdown and 24-hour mortality. Nets not meeting efficacy criteria in cone tests were subsequently evaluated using tunnel tests. Olyset^®^ Plus, a pyrethroid-PBO net, was also tested with the KISUMU strain in cone assays to assess permethrin efficacy, while the added PBO component was evaluated in tunnel tests using unfed 5- to 8-day-old pyrethroid-resistant *An. coluzzii* (AKRON strain), with 24-hour mortality as the key outcome. Interceptor^®^ G2, combining alpha-cypermethrin and chlorfenapyr, was assessed through tunnel tests using 5- to 8-day-old unfed *An. gambiae* (VKPER strain). Efficacy was determined based on 72-hour mortality to capture the delayed killing effect of chlorfenapyr and blood-feeding inhibition. For Royal Guard^®^, which combines alpha-cypermethrin and pyriproxyfen, cone bioassays were conducted with the KISUMU strain to assess the pyrethroid component, and nets failing cone criteria were retested in tunnel assays. The impact of pyriproxyfen on mosquito fertility was assessed through cone bioassays using blood-fed 3- to 5-day-old *An. coluzzii* (AKRON strain), followed by ovary dissections to measure reductions in egg development and fertility.

**Table 2.** Summary of bioassay methods, mosquito strains, and outcome measures used to evaluate ITN bioefficacy by net type and active ingredient

| ITN type | Brand | Active ingredient | Strain | Primary Test Method | Key outcome measures | Remarks/additional tests |
| --- | --- | --- | --- | --- | --- | --- |
| Pyrethroid-only | MAGNet®, Olyset®, DuraNet® | Alpha-cypermethrin | KISUMU-S | Cone bioassays | Knockdown, 24h Mortality | Tunnels for failed nets |
| Pyrethroid-only | Olyset® Net | Permethrin | KISUMU-S | Cone bioassays | Knockdown, 24h Mortality | Tunnels for failed nets |
| Pyrethroid-PBO | Olyset® Plus | Permethrin | KISUMU-S | Cone bioassays | Knockdown, 24h Mortality | Tunnels for failed nets |
|  |  | PBO | AKRON-R | Tunnel tests | 24h Mortality | - |
| Pyrethroid-chlorfenapyr | Interceptor® G2* | chlorfenapyr | VKPER-R | Tunnel tests | 72h Mortality, BFI (alpha) | - |
| Pyrethroid-pyriproxyfen | Royal Guard® | Alpha-cypermethrin | KISUMU-S | Cone bioassays | Knockdown, Mortality | Tunnels for failed nets |
|  |  | Pyriproxyfen | Blood-fed AKRON-R | Cone bioassays | Reduction in Fertility (dissection) | >30% fertility in control Assumed 50% cut off |

\*Interceptor® G2 nets were tested in hut trials prior to bioassays

Bioefficacy was defined as the proportion of ITNs meeting WHO-established criteria i.e knockdown ≥95% or 24/72-hour mortality ≥80% in cone bioassays, and either mortality ≥80% or blood-feeding inhibition ≥90% in tunnel tests. For pyriproxyfen, efficacy was assessed based on a reduction in fertility of ≥50% compared to a control group. Fertility tests were considered valid only if at least 30% of mosquitoes in the control group were fertile. All bioassays were conducted under controlled conditions at 27 ± 2 °C and 75% ± 10% relative humidity.

### Mosquito strains for laboratory bioassays

Cone bioassays and tunnel tests were conducted to monitor the bioefficacy of each active ingredient in the different ITN brands using laboratory-reared susceptible and pyrethroid-resistant strains of *Anopheles gambiae* s.l. All mosquito strains were maintained at the CREC/LSHTM insectary in Cotonou, Benin. The characteristics of each strain are described below.

- KISUMU strain: An insecticide-susceptible *An. gambiae* sensu stricto (*s.s*.) reference strain originating from the Kisumu area in Kenya and was colonized at the CREC/LSHTM insectary.
- AKRON strain: A pyrethroid and carbamate-resistant strain of *Anopheles coluzzii* AKRON originating from Akron (9° 19′ N2° 18′ E), Southern Benin, and maintained at CREC/LSHTM insectary. Resistance is mediated by target site mutations (L1014F *kdr* and *Ace-1R*) and overexpressed cytochrome P450 enzymes [26].
- VKPER strain: A moderately pyrethroid-resistant strain of *An. gambiae* s.s, which originated from the Kou Valley in Burkina Faso. Pyrethroid resistance is mediated mostly by high frequencies of the *kdr* mutation (L1014F).

WHO susceptibility bioassays were conducted during each round of testing to confirm the susceptibility status of each mosquito strain to the insecticides used on the nets. At each time point, approximately 100 mosquitoes from each strain were exposed in batches of 25 using WHO bottle bioassays treated with alpha-cypermethrin (12.5 µg/bottle), chlorfenapyr (100 µg/bottle), and pyriproxyfen (100 µg/bottle), and using WHO tube tests with permethrin (0.75% impregnated papers). For alpha-cypermethrin, chlorfenapyr, and permethrin exposures, unfed mosquitoes aged 2–5 days were exposed for 1 hour, with mortality recorded 24 hours post-exposure for alpha-cypermethrin and permethrin, and 72 hours post-exposure for chlorfenapyr. For pyriproxyfen, blood-fed mosquitoes aged 5–8 days were exposed for 1 hour, and the ovaries of survivors were dissected three days later to assess the proportional reduction in fertility relative to unexposed control mosquitoes, as previously described [21].

In addition, PBO pre-exposure bioassays were performed to assess the involvement of metabolic resistance mechanisms. For these assays, mosquitoes were first exposed to PBO (4%) for 1 hour in WHO bottles or WHO tubes, followed immediately by exposure for 1 hour to either alpha-cypermethrin (12.5 µg/bottle) or permethrin (0.75% impregnated papers), depending on the insecticide tested. Mortality was recorded 24 hours after the final insecticide exposure.

All bioassays were conducted under controlled insectary conditions at 27 °C ± 2 °C and 75% ± 10% relative humidity.

### Experimental hut trial

To complement the laboratory bioassay data, experimental hut trials were conducted using Interceptor^®^ G2 nets withdrawn from households in Mozambique at 6 and 24 months post-distribution. These trials aimed to evaluate the bioefficacy of the product against wild, free-flying mosquitoes under semi-field conditions, particularly considering the unique mode of action of chlorfenapyr. The studies were carried out at the CREC/LSHTM experimental hut station in Covè (7°14′N, 2°18′E), southern Benin, where the local *Anopheles gambiae* s.l. population is characterized by high levels of resistance to pyrethroids and organochlorines, but remains susceptible to other insecticide classes, including chlorfenapyr. The vector population consists of a mix of *An. coluzzii* and *An. gambiae s.s.*, with resistance primarily driven by the L1014F knockdown resistance (kdr) mutation and elevated expression of cytochrome P450 enzymes [22, 23].

Field-aged Interceptor^®^ G2 nets were evaluated alongside new, unused Interceptor^®^ G2 nets and pyrethroid-only ITNs. Five treatments were compared across two trials; one conducted with Interceptor^®^ G2 nets aged 6 months in the field and another with nets aged 24 months.

1. Untreated net
2. MagNet^®^ (new unwashed)
3. Interceptor^®^ (new unwashed)
4. Interceptor^®^ G2 (new unwashed)
5. Interceptor^®^ G2 – field collected from Mozambique at 6 or 24 months

#### Hut trial design

Treatments were rotated weekly across the five experimental huts using a randomized Latin Square Design to minimize potential bias due to hut position. Data collection was conducted over a 30-night period spanning five weeks, with six consecutive nights of collection each week. The seventh day was reserved for cleaning and airing the huts in preparation for the next rotation. For the field-collected Interceptor^®^ G2 nets, 30 replicate nets were tested per treatment, with nets rotated daily. For other treatments, six replicate nets were used, with each net tested every night of the week. New nets were given six 4x4cm deliberate holes on each panel in line with WHO guidelines, to allow assessment of blood-feeding protection.

Each night from 21:00 to 06:00, five consenting human volunteers slept in the huts to attract wild, free-flying mosquitoes. Each morning, volunteers collected mosquitoes from hut compartments (under the net, inside the room, and in the veranda) using a torch and aspirator, and placed them in labeled plastic cups. These collections were transferred to the field laboratory for morphological identification and assessed for immediate mortality and blood-feeding status. Surviving female *An gambiae* s.l. were maintained at 27 ± 2°C and 75 ± 10% relative humidity with access to a 10% glucose solution. Delayed mortality was recorded after 72 hours.

The primary outcome measures used to express the efficacy of the experimental hut treatments against wild pyrethroid-resistant *An. gambiae* s.l. entering the huts and compare the impact of the Interceptor^®^ G2 to the pyrethroid-only net were:

- **Mortality (%)**: Proportion of mosquitoes that died within 72 hours post-collection.
- **Blood-feeding inhibition (%)**: Reduction in blood-feeding in treated huts relative to the control

Other secondary outcome measures were:

- **Entry rate**: Total number of mosquitoes collected per hut.
- **Deterrence (%)**: Reduction in mosquito entry in treated huts compared to the untreated control hut.
- **Exophily (%)**: Proportion of mosquitoes found in the veranda, indicating treatment-induced exiting behavior.
- **Inside net (%)**: Proportion of mosquitoes collected inside the net.
- **Blood-feeding rate (%)**: Proportion of mosquitoes that were blood-fed.

### Chemical analysis methods

Approximately 600 net pieces, each measuring 30x30 cm, that were obtained from positions on the ITN adjacent to those used for bioassays were preserved for chemical content analysis at each timepoint. The samples were wrapped in aluminum foil and stored at 4°C (±2°C) before being shipped to the Centre Wallon de Recherches Agronomiques (CRA-W) in Belgium for active ingredient (AI) content testing. The methods for AI extraction have been previously described [9, 24]. Gas Chromatography with Flame Ionisation Detection (GC-FID) was used to quantify the AI content. To obtain a single chemical content result per AI for each ITN type sampled at each timepoint, net pieces from the same net were pooled.

### Statistical analysis

For each ITN and each timepoint entomological outcomes in bioassays (knockdown, mortality, blood-feeding, passage etc) were pooled across the different ITN pieces tested. A chi-squared test was used to assess the proportion of nets of each ITN type passing the WHO criteria for pyrethroids, chlorfenapyr and pyriproxyfen bioefficacy based on combined cone and tunnel tests. For experimental hut trial data, proportional outcomes (72 h mortality, blood-feeding, exophily) were analysed using logistic regression, while differences in count outcomes (entry) were analysed using negative binomial regression. Each outcome was modelled separately, with adjustments for variation between huts, sleepers, trial timepoint and trial days included as fixed effects.

#### Ethical approval and consent to participate

Ethical approval for withdrawal of the field-aged nets was obtained from the Comité Nacional de Bioética para a Saúde em Moçambique (Ref:/424/CNBS/20), while the ethics review boards of the Ministry of Health in Benin gave ethical approval for the hut trial. Informed written consent was obtained from all human volunteers before their participation. A stand-by nurse was available throughout the trials to assess any volunteers presenting with febrile symptoms or an adverse reaction to the test items. The methods described in this paper followed relevant guidelines and regulations. Approval for using guinea pigs for tunnel tests was granted by LSHTM Animal Welfare Ethics Review Board (AWERB) (2020-01B).

## Results

### Susceptibility of test strains

The susceptibility bioassays confirmed that the KISUMU strain was fully susceptible to all insecticides used on the study nets (Figure 2). In contrast, the pyrethroid-resistant VKPER and AKRON strains showed resistance to pyrethroids, with mortality rates of <60% following exposure to alphacypermethrin (12.5 µg) and permethrin (0.75%). Both resistant strains, however, remained fully susceptible to chlorfenapyr (100 µg) and pyriproxyfen, with responses exceeding 95%. Pre-exposure to PBO followed by permethrin (0.05%) substantially restored activity against the AKRON strain (from 19% to 77%), but only modestly against the VKPER strain (from 57% to 68%). These results confirm a stronger role of metabolic enzymes in pyrethroid resistance in the AKRON strain, highlighting its suitability for evaluating pyrethroid–PBO ITNs. Overall, the findings validate the use of these different test strains for assessing ITN insecticides according to their resistance profiles.

**Figure 2:**
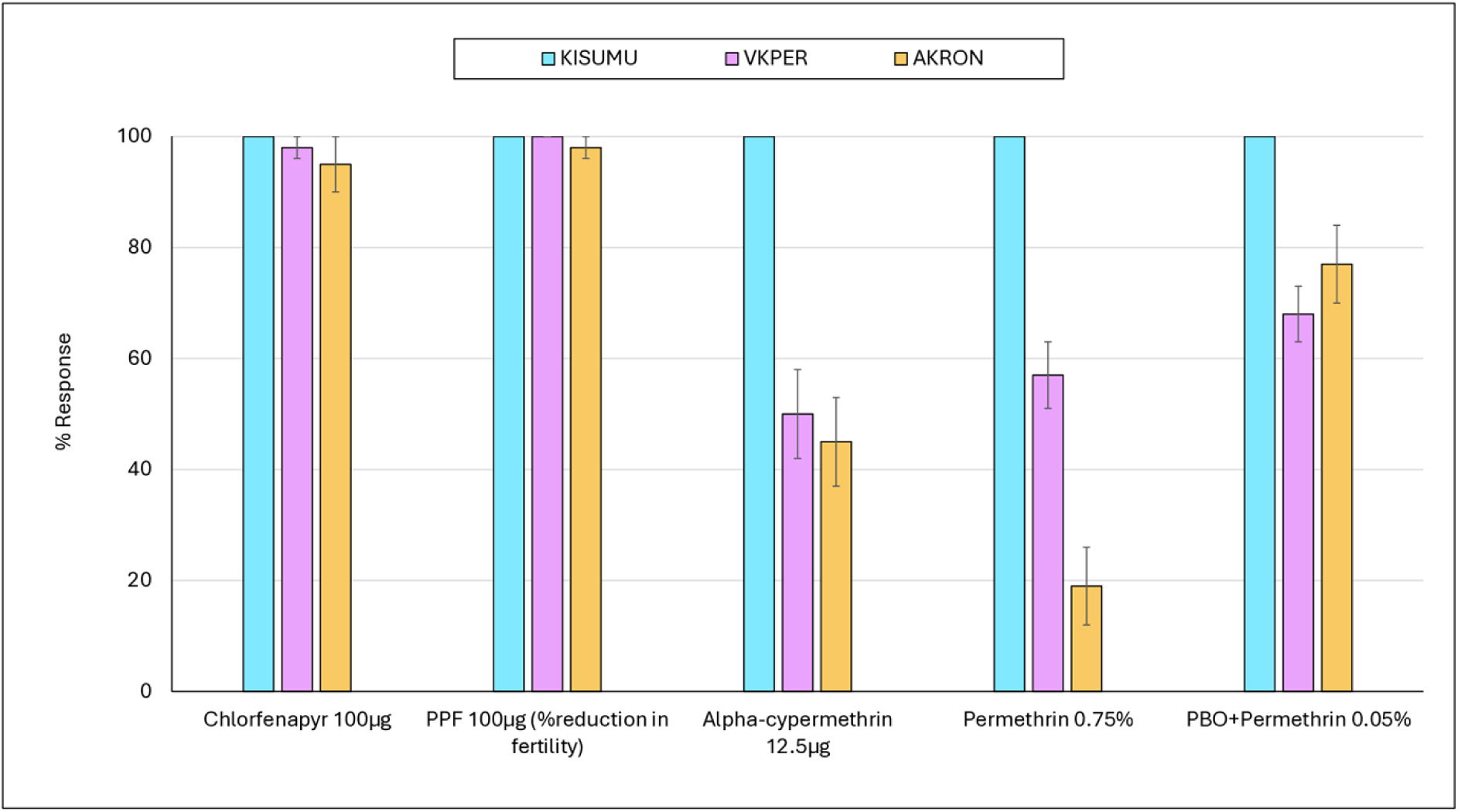
Susceptibility of test strains mosquitoes (KISUMU, VKPER and AKRON) to insecticides applied on study nets. Approximately 100 mosquitoes were exposed to each insecticide at each time point. Susceptibility to pyriproxyfen was assessed by dissecting mosquito ovaries post exposure to determine reduction in fertility. PPF=pyriproxyfen.

### Bioefficacy results for pyrethroid-only ITNs

A total of 10,783 *Anopheles gambiae* (KISUMU strain) mosquitoes were exposed to pyrethroid-only ITNs in WHO cone bioassays conducted at baseline and at 6, 12, and 24 months post-distribution (Table 3). Mortality remained consistently high, with mean values of 88.7% at baseline, 87.1% at 6 months, 98.3% at 12 months, and 99.4% at 24 months. Knockdown rates also exceeded 87% at all time points, reaching nearly 100% at 12 and 24 months. At 6 and 24 months, a small number of net pieces (7 and 1, respectively) that failed to meet cone bioassay criteria were subjected to tunnel tests. In these tests, mosquito mortality remained high (99.4% and 87.3%), and blood-feeding inhibition was 99.3% and 90.3%, respectively. Overall, all ITNs tested at each time point (38/38 at baseline and 30/30 at 6, 12, and 24 months) met WHO bioefficacy criteria, either through cone bioassays alone or when supplemented by tunnel tests. These results confirm the sustained bioefficacy of pyrethroid-only ITNs up to 24 months after distribution.

**Table 3.** Bioefficacy of pyrethroid-only nets tested against the susceptible *Anopheles gambiae* (KISUMU) strain in cone bioassays and tunnel tests at baseline, 6, 12, and 24 months post-distribution in Mozambique. *Only nets failing WHO criteria in cone bioassays were subjected to tunnel tests*.

|  | Baseline | 6 months | 12 months | 24 months |
| --- | --- | --- | --- | --- |
| *Cone bioassays* |  |  |  |  |
| N ITNs tested | 38 | 30 | 30 | 30 |
| N pieces tested | 190 | 150 | 150 | 150 |
| N exposed | 2913 | 1504 | 3252 | 3114 |
| N Knockdown | 2781 | 1313 | 3250 | 3080 |
| N dead | 2585 | 1144 | 3194 | 3062 |
| % Knockdown | 95.5 | 87.3 | 99.9 | 98.9 |
| % dead | 88.7 | 87.1 | 98.3 | 99.4 |
| *Tunnel tests with failed net pieces* |  |  |  |  |
| N pieces tested | 0 | 7 | 0 | 1 |
| N exposed | n/a | 658 | n/a | 102 |
| N passage | n/a | 239 | n/a | 45 |
| N dead | n/a | 654 | n/a | 89 |
| N blood-fed | n/a | 4 | n/a | 8 |
| % Passage | n/a | 36.3 | n/a | 44.1 |
| % dead | n/a | 99.4 | n/a | 87.3 |
| % blood-fed | n/a | 0.6 | n/a | 7.8 |
| % BFI | n/a | 99.3 | n/a | 90.3 |
| N ITNs passing | 38/38 | 30/30 | 30/30 | 30/30 |

### Bioefficacy results for Pyrethroid-PBO ITNs (Olyset^®^ Plus)

#### Permerthin bioefficacy in Olyset^®^ Plus

A total of 9,369 *Anopheles gambiae* (KISUMU) mosquitoes were exposed to Olyset^®^ Plus nets in cone bioassays to evaluate the bioefficacy of the permethrin component at baseline, 6, 12, and 24 months post-distribution (Table 4). At baseline, knockdown and mortality rates were 100% and 99.4%, respectively. Knockdown remained consistently high over time (>90%), but mortality declined progressively from 71.4% at 6 months to 76.3% at 12 months and 59.1% at 24 months. Nets that failed to meet WHO criteria in cone bioassays were further tested in tunnel assays, with 9, 6, and 14 pieces assessed at 6, 12, and 24 months, respectively. In tunnel tests, mosquito mortality remained high (≥96.7%), and blood-feeding inhibition was consistently above 95%. Despite reduced mortality in cone tests over time, all nets tested at each time point, met WHO bioefficacy criteria when cone and tunnel results were combined.

**Table 4.**
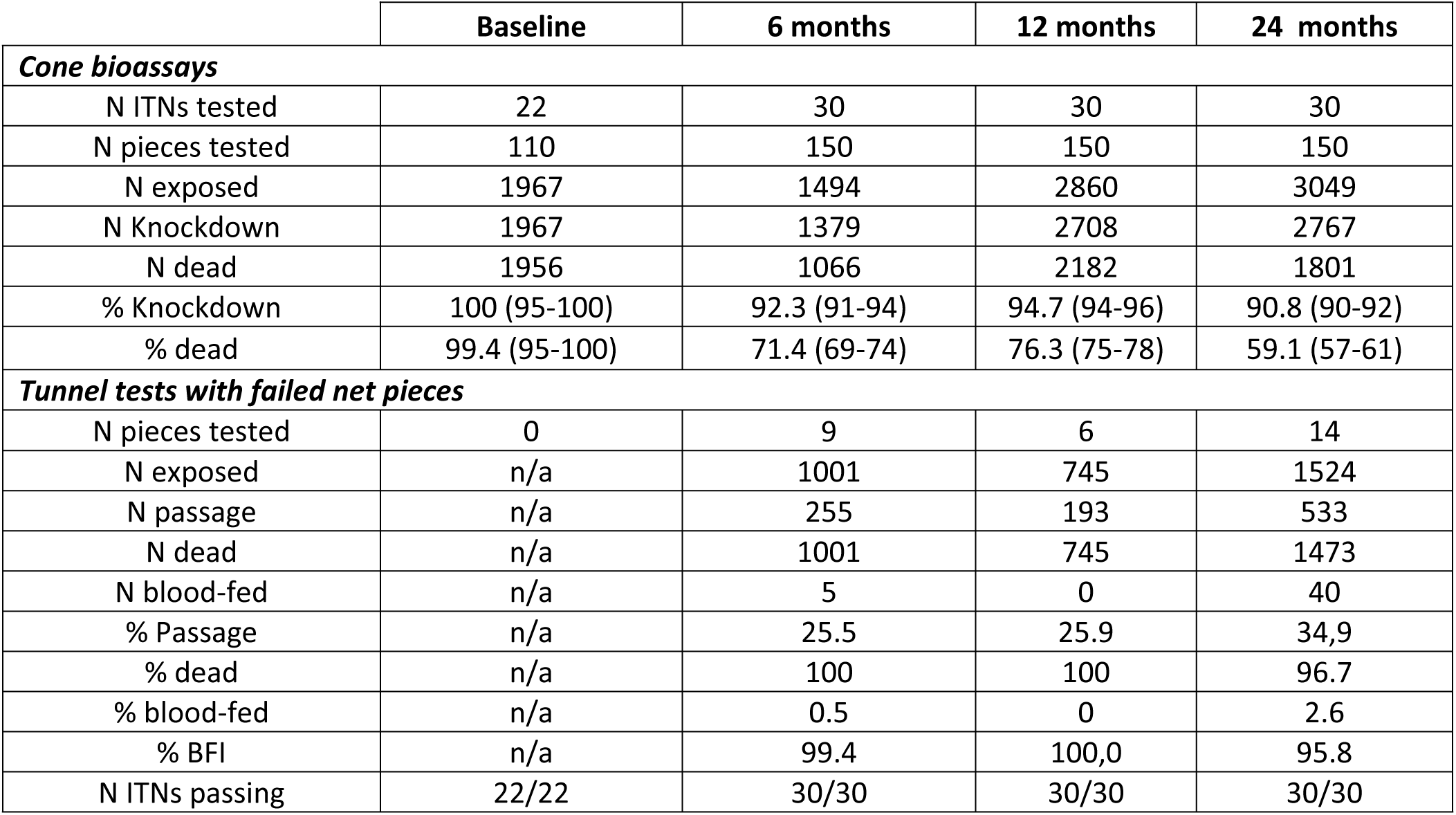
Bioefficacy of the permethrin component in Olyset^®^ Plus nets assessed using cone and tunnel bioassays with *Anopheles gambiae* (KISUMU) at baseline, 6, 12, and 24 months post-distribution in Mozambique. *Only nets failing WHO criteria in cone bioassays were subjected to tunnel tests*.

#### PBO bioefficacy in Olyset^®^ Plus

A total of 11,211 pyrethroid-resistant *Anopheles coluzzii* (AKRON strain) mosquitoes were exposed to Olyset^®^ Plus nets in tunnel tests to evaluate the bioefficacy of the PBO component at baseline, 6, 12, and 24 months post-distribution (Table 5). At baseline, the nets demonstrated high efficacy, with 98.5% mosquito mortality and 98.8% blood-feeding inhibition (BFI), resulting in all nets (16/16) meeting WHO criteria. However, efficacy declined steadily over time, with mortality dropping to 81.4% at 6 months, 76.5% at 12 months, and 50.9% at 24 months. Despite BFI remaining relatively high (≥74.8%) throughout, the proportion of nets meeting WHO bioefficacy thresholds declined to 22/30 at 6 months, 16/30 at 12 months, and 14/30 at 24 months. These results demonstrate a progressive reduction in the efficacy of the PBO component of Olyset^®^ Plus against pyrethroid-resistant mosquitoes over two years of field use in Mozambique.

**Table 5.** Bioefficacy of the PBO component in Olyset^®^ Plus nets tested against pyrethroid-resistant *Anopheles coluzzii* (AKRON strain) in tunnel assays at baseline, 6, 12, and 24 months post-distribution in Mozambique.

|  | Baseline | 6 months | 12 months | 24 months |
| --- | --- | --- | --- | --- |
| N ITNs tested | 16 | 30 | 30 | 30 |
| N pieces tested | 16 | 30 | 30 | 30 |
| N exposed | 1637 | 3163 | 3350 | 3061 |
| N passage | 741 | 1765 | 1755 | 1350 |
| N dead | 1612 | 2574 | 2561 | 1559 |
| N blood-fed | 18 | 663 | 514 | 414 |
| % Passage | 45.3 | 55.8 | 52.4 | 44.1 |
| % dead | 98.5 | 81.4 | 76.5 | 50.9 |
| % blood-fed | 1.1 | 21 | 15.3 | 13.5 |
| % BFI | 98.8 | 74.8 | 81 | 80.6 |
| N ITNs passing | 16/16 | 22/30 | 16/30 | 14/30 |

### Bioefficacy results of Pyrethroid-pyriproxyfen ITNs (Royal Guard)

Royal Guard^®^ demonstrated consistently high alpha-cypermethrin bioefficacy over 24 months, with cone bioassays using the susceptible KISUMU strain showing knockdown rates of ≥99.6% and mortality rates of ≥99.5% at all time points (Table 6). All nets tested (100%) met WHO bioefficacy criteria at baseline (19/19), 6 months (30/30), 12 months (30/30), and 24 months (31/31). In contrast, the pyriproxyfen component showed a decline in fertility reduction over time when tested against blood-fed pyrethroid-resistant AKRON mosquitoes. At baseline, 100% fertility reduction was observed (0/92 fertile mosquitoes), but this decreased to 46% at 6 months, 53.5% at 12 months, and 72.3% at 24 months. Consequently, the proportion of ITNs meeting pre-defined thresholds for pyriproxyfen-induced sterilization declined at 6 months relative to baseline (21/21 vs 13/30) and eventually increased at subsequent timepoints; 18/30 at 12 months, and 27/31 at 24 months.

**Table 6.** Bioefficacy of the pyrethroid and pyriproxyfen components in Royal Guard^®^ nets tested against susceptible *Anopheles gambiae* (KISUMU strain) and pyrethroid-resistant *Anopheles coluzzii* (AKRON strain) in cone bioassays at baseline, 6, 12, and 24 months post-distribution

|  | Baseline | 6 months | 12 months | 24 months |
| --- | --- | --- | --- | --- |
| *Cone bioassays with KISUMU to investigate alpha-cypermethrin bioefficacy* |  |  |  |  |
| N ITNs tested | 19 | 30 | 30 | 31 |
| N pieces tested | 95 | 150 | 150 | 155 |
| N exposed | 1638 | 1457 | 3106 | 3170 |
| N Knockdown | 1638 | 1451 | 3106 | 3164 |
| N dead | 1638 | 1449 | 3100 | 3156 |
| % Knockdown | 100 | 99.6 | 100 | 99.8 |
| % dead | 99.6 | 99.5 | 99.8 | 99.6 |
| N ITNs passing | 19/19 | 30/30 | 30/30 | 31/31 |
| *Cone bioassays with blood-fed AKRON to investigate pyriproxyfen bioefficacy* |  |  |  |  |
|  | Baseline | 6 months | 12 months | 24 months |
| N ITNs tested | 21 | 30 | 30 | 31 |
| N pieces tested | 105 | 150 | 150 | 155 |
| N exposed | 1842 | 1404 | 3010 | 3508 |
| N dissected | 92 | 278 | 723 | 715 |
| N fertile | 0 | 150 | 329 | 193 |
| N infertile | 92 | 128 | 394 | 522 |
| % Fertile | 100 | 54 | 45.5 | 26,9 |
| % Reduction in Fertility | 100 | 46 | 53.5 | 72.3 |
| N ITNs passing | 21/21 | 13/30 | 18/30 | 27/31 |

### Bioefficacy results of Pyrethroid-chlorfenapyr ITNs (Interceptor^®^ G2)

A total of 10,845 pyrethroid-resistant *Anopheles gambiae* VKPER mosquitoes were exposed to Interceptor^®^ G2 nets in tunnel tests over the 24-month evaluation period to assess the durability of the bioefficacy of the chlorfenapyr component (Table 7). Mosquito mortality was high at baseline (96.6%) and remained above 90% throughout. Blood-feeding inhibition improved from 77.0% at baseline to 93.6% at 12 months but dropped significantly to 46.3% by 24 months. All nets tested at baseline and 6 months passed WHO efficacy thresholds and this remained high at 12 months (29/30) and 24 months (28/30).

**Table 7.** Bioefficacy of Interceptor^®^ G2 nets tested in tunnel assays against pyrethroid-resistant *Anopheles gambiae* (VKPER strain) at baseline, 6, 12, and 24 months post-distribution

|  | Baseline | 6 months | 12 months | 24 months |
| --- | --- | --- | --- | --- |
| N ITNs tested | 16 | 30 | 30 | 30 |
| N pieces tested | 16 | 30 | 30 | 30 |
| N exposed | 1584 | 3006 | 3107 | 3148 |
| N passage | 630 | 1434 | 1381 | 1548 |
| N dead | 1530 | 2902 | 2979 | 2858 |
| N blood-fed | 169 | 368 | 172 | 351 |
| % Passage | 39.8 | 47.7 | 44.4 | 49.2 |
| % dead | 96.6 | 96.5 | 95.8 | 90.8 |
| % blood-fed | 10.7 | 12.2 | 5.5 | 49.2 |
| % BFI | 77.0 | 85.3 | 93.6 | 46.3 |
| N ITNs passing | 16/16 | 30/30 | 29/30 | 28/30 |

### Overall ITN pass rates in laboratory bioassays

The percentages of ITNs passing WHO bioefficacy criteria for each active ingredient when new and unused and at 6, 12, and 24 months are presented in Figure 3. Pyrethroid components in the pyrethroid-only nets (Olyset^®^ Net, MAGNet^®^, and DuraNet^®^), Olyset^®^ Plus (permethrin), and Royal Guard^®^ (alpha-cypermethrin) consistently achieved 100% pass rates in cone bioassays with the susceptible *An gambiae ss* KISUMU strain across all time points. In contrast, the efficacy of the PBO component in Olyset^®^ Plus declined steadily over time, with pass rates dropping from approximately 73% at 6 months to 50% at 12 months and 40% at 24 months. For Royal Guard^®^, while alpha-cypermethrin maintained 100% efficacy throughout, the pyriproxyfen component showed an unusual upward trend in performance, with pass rates increasing from 43% at 6 months to 60% at 12 months and 87% at 24 months. Interceptor^®^ G2, also demonstrated strong and sustained efficacy for chlorfenapyr in tunnel tests with the pyrethroid-resistant VKPER strain, maintaining ITN pass rates of 92%-100% over the 24-month period.

**Figure 3.**
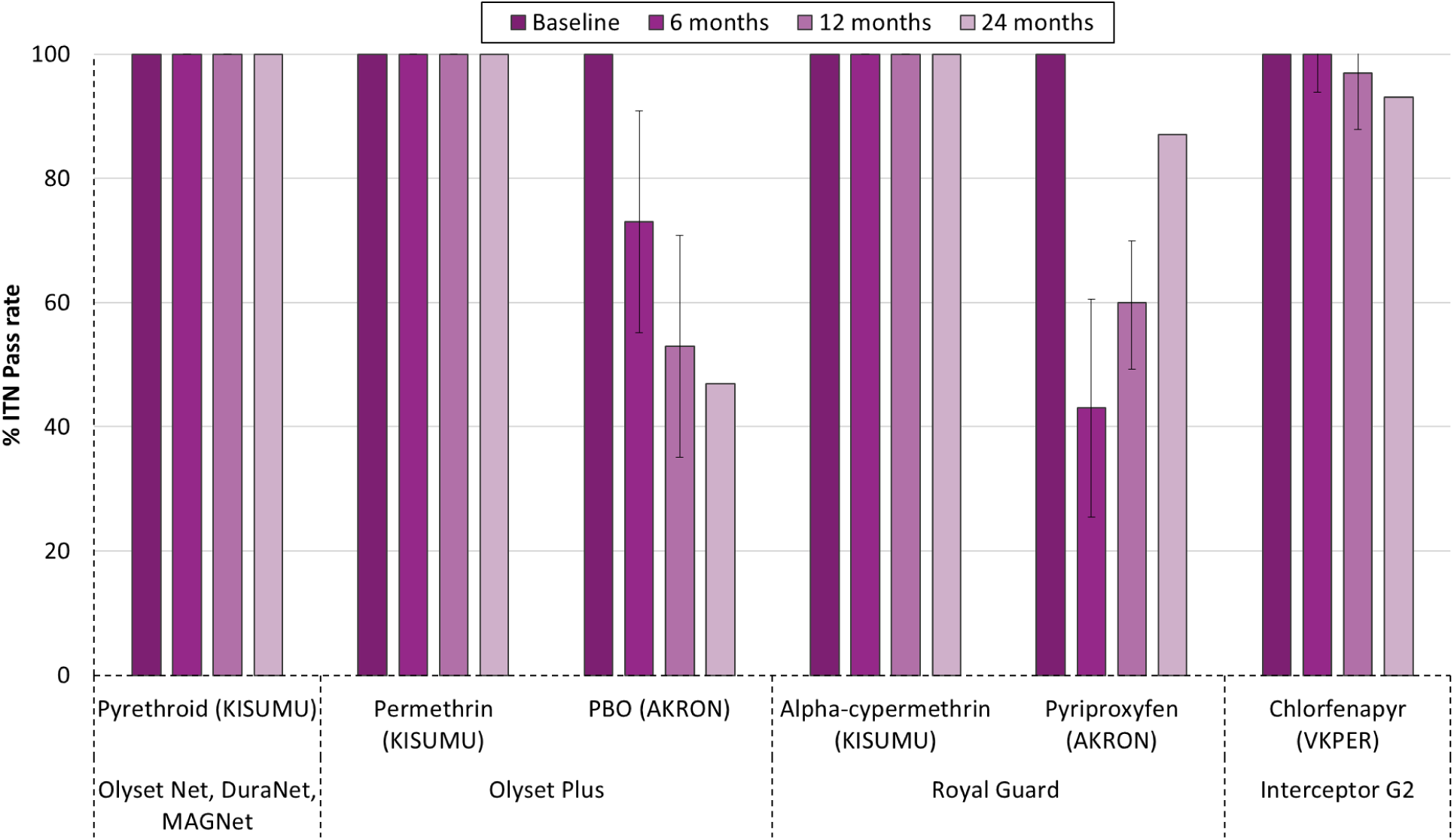
Proportion of ITNs passing WHO bioefficacy criteria by active ingredient and time point. The figure shows the percentage of ITNs that met WHO bioefficacy criteria at baseline (new and unused) and at 6, 12 and 24 months after distribution, stratified by active ingredient and mosquito strain used in bioassays. Pyrethroid-only ITNs (Olyset^®^ Net, DuraNet^®^, MAGNet) and the alpha-cypermethrin component of Royal Guard^®^ consistently achieved 100% pass rates using the *Anopheles gambiae* KISUMU strain. Efficacy of the PBO component in Olyset^®^ Plus declined over time, while the pyriproxyfen component in Royal Guard^®^ showed an increasing trend. Interceptor^®^ G2, evaluated using the VKPER pyrethroid-resistant strain, maintained high pass rates with its chlorfenapyr component. Error bars represent 95% confidence intervals.

### Experimental hut results

A total of 2,326 wild, free-flying pyrethroid-resistant *An gambiae* s.l. mosquitoes were collected in experimental huts in Cove, Benin during the trials with new and field-aged Interceptor^®^ G2 nets collected from households in Mozambique at 6 and 24 months post-distribution. Results are presented in Figure 4 for mortality and Figure 5 for blood-feeding inhibition with further details available in Table S1.

**Figure 4.**
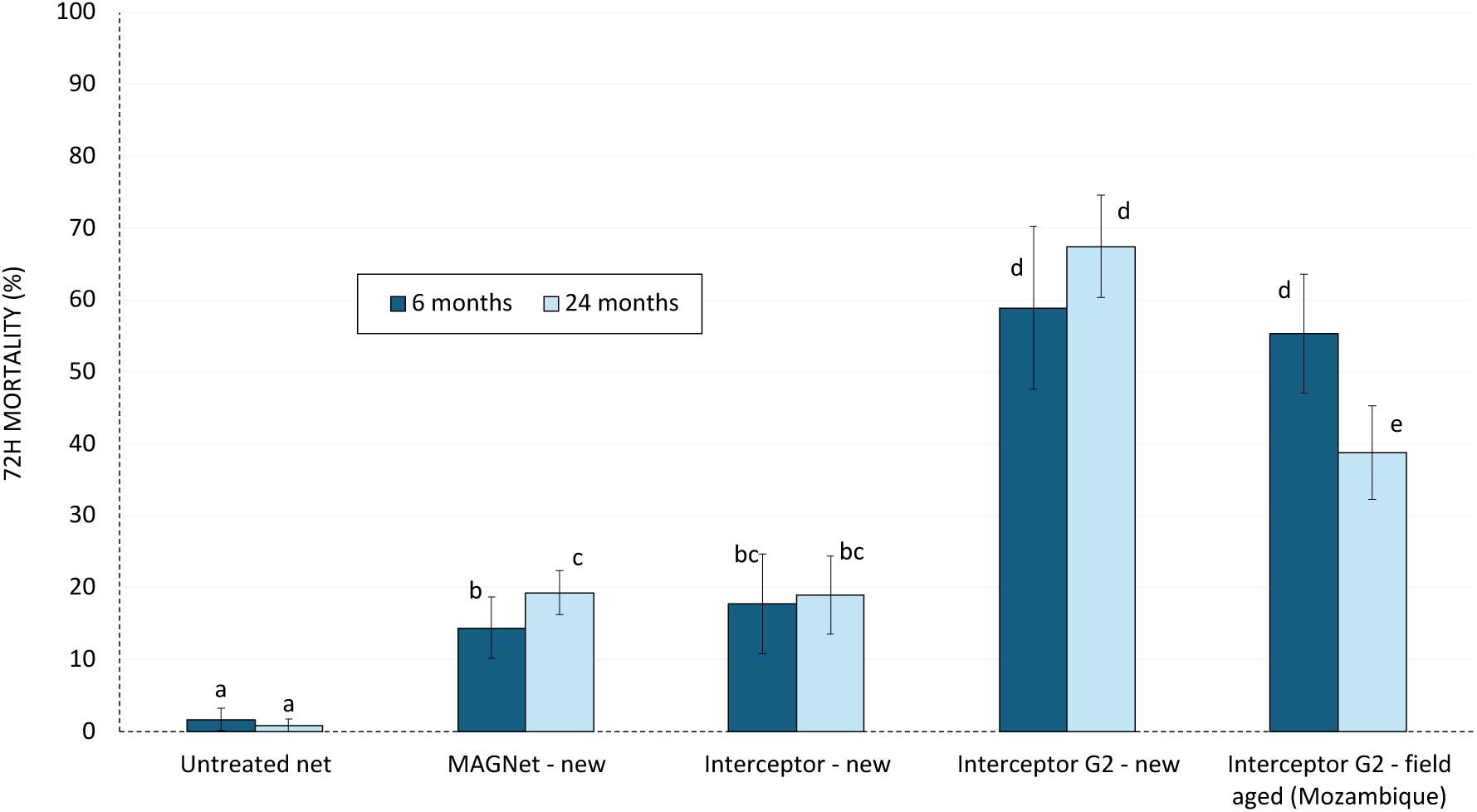
Mortality (72 h) of wild free-flying pyrethroid-resistant *An gambiae s.l.* in experimental hut trials in Cove Benin with new and field-aged Interceptor^®^ G2 nets collected from Mozambique at 6 and 24 months post-distribution. *Two separate hut trials were performed at two timepoints; one with 6 months old net field aged Interceptor^®^ G2 nets and another at 24 months old field aged Interceptor^®^ G2 nets*.

**Figure 5.**
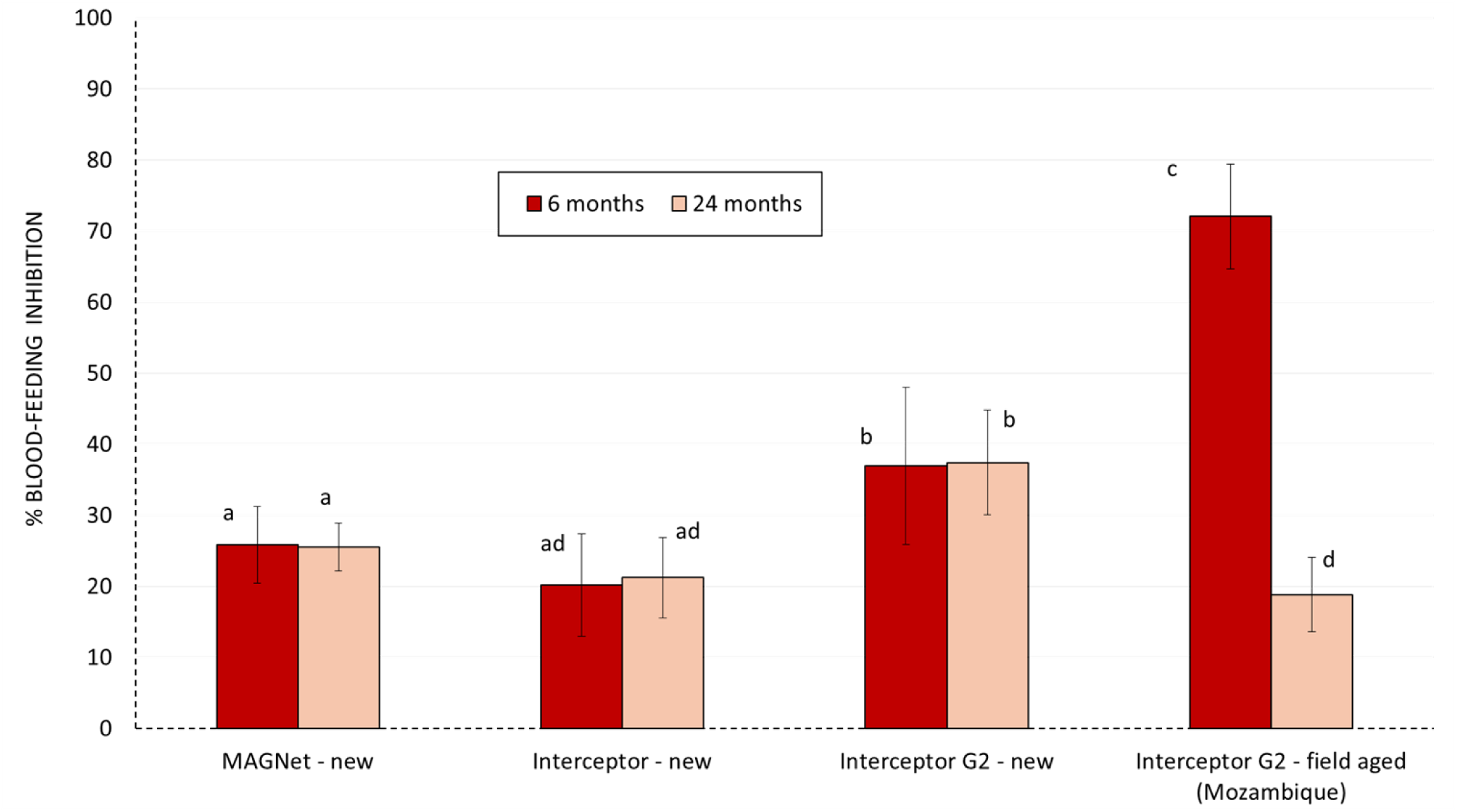
Blood-feeding inhibition of wild free-flying pyrethroid-resistant *An gambiae* sl in experimental hut trials in Cove Benin with new and field-aged Interceptor^®^ G2 nets collected from Mozambique at 6 and 24 months post-distribution. *Two separate hut trials were performed at two timepoints; one with 6 months old net field aged Interceptor^®^ G2 nets and another at 24 months old field aged Interceptor^®^ G2 nets*.

#### Mortality rates

Mortality with untreated nets remained consistently low (<5%) at both time points. The new pyrethroid-only nets (MAGNet^®^ and Interceptor^®^) induced moderate mortality (15–20%) that did not differ substantially between both trials (∼19%), reflecting their limited efficacy against the pyrethroid-resistant Cove vector population. In contrast, new Interceptor^®^ G2 nets achieved significantly higher mortality at both trial timepoints (59% at 6 months and 67% at 24 months; p < 0.05), demonstrating the enhanced killing effect provided by chlorfenapyr. Field-aged Interceptor^®^ G2 nets from Mozambique showed reduced performance over time, with mortality decreasing from 55% at 6 months to 39% at 24 months (p = 0.001). While still more effective than pyrethroid-only nets, the 24-month field-aged Interceptor^®^ G2 nets were significantly less effective than the new Interceptor^®^ G2 nets tested at the same time point (p < 0.05), indicating a decline in bioefficacy with operational use and ageing.

#### Blood-feeding inhibition rates

Blood-feeding inhibition with new pyrethroid-only nets (MAGNet^®^ and Interceptor^®^) was low (20–30%) and did not vary significantly between time points, indicating limited personal protection against pyrethroid-resistant *An. gambiae s.l.*. New Interceptor^®^ G2 nets provided moderate and consistent levels of blood-feeding inhibition (approximately 37% at both time points), with no substantial decline observed across both trials. In contrast, field-aged Interceptor^®^ G2 nets showed a marked reduction in blood-feeding inhibition with age: 72% at 6 months versus only 19% at 24 months (p < 0.05). The high inhibition observed at 6 months suggests strong initial personal protection, while the significant drop by 24 months indicates a loss of this protective effect over time.

### Chemical analysis results

At baseline, all ITNs had full active ingredient content (100%), consistent with manufacturer specifications (Figure 6). Over time, the rate of active ingredient loss varied across products and insecticide types. Pyrethroid-only nets such as Olyset^®^ and DuraNet^®^ maintained relatively high levels of permethrin and alpha-cypermethrin (≥80%) after 24 months. In contrast, MAGNet^®^ exhibited a more pronounced decline, retaining only 73% of alpha-cypermethrin at 12 months and 56% at 24 months. Olyset^®^ Plus showed a sharp drop in PBO content, retaining just 48% at 12 months and only 8% at 24 months, while permethrin levels declined more gradually to 73% and 48%, respectively. For Royal Guard^®^, alpha-cypermethrin and pyriproxyfen content declined steadily, with both active ingredients retaining approximately 47–67% by 24 months. Interceptor^®^ G2 nets retained alpha-cypermethrin at 66% and chlorfenapyr at just 32% by 24 months, suggesting a more rapid loss of chlorfenapyr than pyrethroid. These findings indicate that while some pyrethroids remained relatively stable over time, synergists and novel actives such as PBO and chlorfenapyr showed greater degradation during operational use.

**Figure 6.**
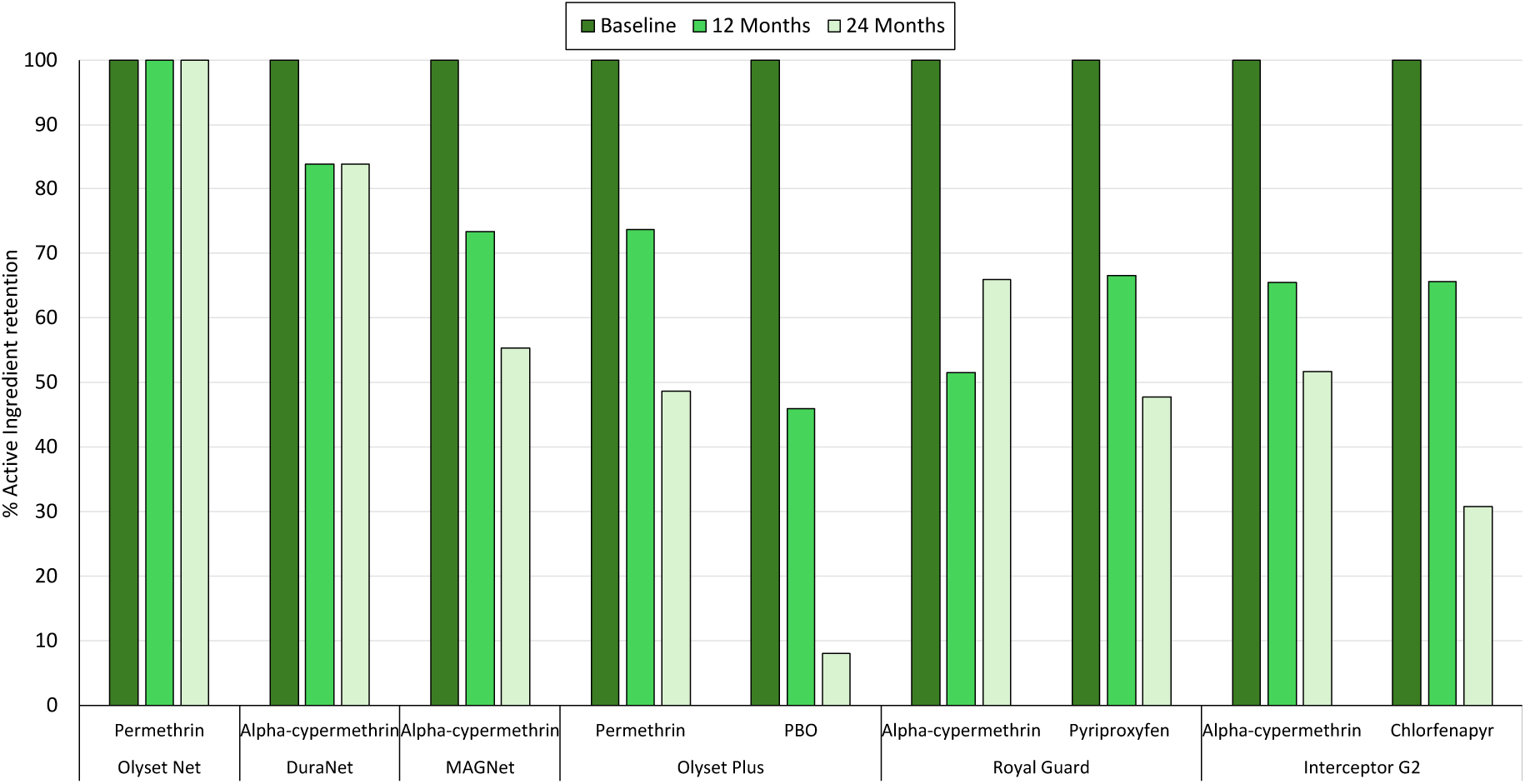
Percentage of active ingredient retention in insecticide-treated nets (ITNs) at baseline, 12 months, and 24 months post-distribution in Mozambique.

## Discussion

This study evaluated the durability of the insecticidal activity and chemical content of next-generation ITNs over 24 months post distribution to householders in communities in Mozambique during the New Nets project (2020–2022). Using both laboratory bioassays and experimental hut trials, our findings demonstrate that while all ITN types retained substantial bioefficacy over 24 months, there were clear differences in the performance of specific active ingredients, particularly among the newer insecticide classes.

The pyrethroid-only ITNs (MAGNet^®^, Olyset^®^ Net, and DuraNet^®^) demonstrated consistently high efficacy in WHO cone and tunnel bioassays with the susceptible strain over the 24-month evaluation period, with 100% of nets meeting WHO criteria at all time points. This was supported by chemical analysis, which showed substantial retention of the pyrethroid insecticide, ranging from 50% to 100% at endline. These results highlight the chemical durability of pyrethroids on ITNs and are consistent with findings from a similar study conducted in Benin [25]. However, despite their chemical persistence, the limited effectiveness of pyrethroid-only ITNs against wild pyrethroid-resistant vector populations is well established [22, 26]. This was further confirmed by the experimental hut trial in the current study, which recorded low mortality rates (<20%) in wild, resistant *An. gambiae* s.l., indicating poor field performance of unused pyrethroid-only ITNs. These findings reinforce the need for next-generation ITNs with additional active ingredients to overcome pyrethroid resistance and sustain vector control impact.

Although Olyset^®^ Plus maintained strong performance in WHO cone and tunnel bioassays using susceptible mosquitoes—highlighting the durability of its pyrethroid component—the efficacy of the PBO component against pyrethroid-resistant mosquitoes declined steadily over time. The proportion of nets meeting WHO bioefficacy thresholds for PBO-specific efficacy dropped from 73% at 6 months to just 40% at 24 months. This decline closely mirrored the rapid loss of PBO content, which fell to only 8% of the original concentration by 24 months. These findings are consistent with earlier reports indicating that, while PBO plays a critical role in enhancing pyrethroid efficacy in areas with metabolic resistance, it is susceptible to rapid degradation under operational conditions [6, 25, 27, 28]. This degradation significantly limits the duration of enhanced protection provided by PBO nets.

Royal Guard^®^, which incorporates both alpha-cypermethrin and pyriproxyfen, showed high and stable pyrethroid efficacy in cone bioassays throughout the study, but variable performance of the pyriproxyfen component. Fertility reduction declined initially at 6 months (46%) but improved again by 24 months (72%), with pass rates increasing from 43% to 87%. The reasons for this unexpected rebound are unclear but may reflect variability in exposure conditions or mosquito physiological status at the time of testing. Despite this, the data suggest that Royal Guard^®^ may offer meaningful sterilizing effects beyond one year of use in the field, supporting its added value over pyrethroid-only ITNs in pyrethroid-resistant areas as indicated in WHO guidelines [5].

Interceptor^®^ G2 consistently induced high mortality in tunnel tests against the pyrethroid-resistant VKPER mosquito strain throughout the 24-month evaluation period showing durability of the improved insecticidal effect of chlorfenapyr in the net. In experimental hut trials, mortality of wild pyrethroid-resistant malaria vectors remained significantly higher with Interceptor^®^ G2 compared to pyrethroid-only nets even at 24 months, corroborating findings from multiple hut trials conducted across Africa [10, 29-31]. Chemical analysis showed that alpha-cypermethrin content remained relatively stable over time, whereas chlorfenapyr levels declined to 32% of baseline by 24 months. This degradation likely contributed to some loss of efficacy; however, the net continued to demonstrate substantially improved killing of wild vectors compared to new pyrethroid-only nets after two years of household use. These results reinforce evidence from cluster randomized controlled trials (cRCTs) across Africa supporting the superior efficacy of Interceptor^®^ G2 over 2 years of community use [16], and providing further justification for WHO’s strong recommendation of this ITN class for malaria vector control in areas of resistance [5].

Although Interceptor^®^ G2 nets sustained relatively high mortality (>90%) in tunnel tests across all timepoints, the hut trial demonstrated a decline in efficacy at 24 months compared to 6 months. While these discrepancies between laboratory bioassays and hut trial outcomes may reflect inherent differences in the mosquito strains tested, the results indicate that experimental hut trials with field-aged nets generate valuable complementary data to laboratory bioassays. Both tunnel tests and hut trials showed a marked decline in blood-feeding inhibition with Interceptor^®^ G2 at 24 months, indicating a reduced ability to provide personal protection as the nets age. This contrasts with findings from hut trials in Benin, where aged Interceptor^®^ G2 nets showed improved blood-feeding inhibition compared to earlier time points [23]. In the present study, however, blood-feeding inhibition was higher with 6-month-old nets than with new nets, possibly reflecting the stabilization of insecticide bioavailability after initial washing or handling. These differences highlight the potential influence of local environmental conditions and household practices such as net maintenance, frequency of use/washing, and sleeping arrangements on ITN performance, and underscore the need to evaluate both mortality and blood-feeding outcomes when assessing their long-term efficacy under experimental hut conditions.

A key limitation of this study is that net durability was evaluated only up to 24 months, rather than the full intended three-year lifespan, due to resource constraints and project timelines. Consequently, the long-term performance of next-generation nets in Mozambique beyond two years remains unclear. Findings from Benin showed that dual active ingredient nets, such as Interceptor^®^ G2, exhibit substantial declines in active ingredient content and bioefficacy by the third year post-distribution, leading to a loss of their improved epidemiological impact over pyrethroid-only nets [25, 32]. To better understand the full operational lifespan and performance trajectory of these nets, extended follow-up studies covering the entire three-year period across varied ecological and transmission settings are recommended. Such studies would offer critical insights to guide replacement intervals and overall decision making for malaria control, including policy development, procurement planning, and implementation strategies.

## Conclusion

This study provides important operational evidence on the durability of next-generation insecticide-treated nets (ITNs) deployed in Mozambique through the New Nets Project, revealing distinct differences in performance across net types and active ingredients over 24 months of household use. Although all ITNs maintained bioefficacy against susceptible mosquito strains, only Interceptor^®^ G2 sustained high efficacy against pyrethroid-resistant vectors throughout the study period, reaffirming the enhanced performance of pyrethroid-chlorfenapyr nets. Nonetheless, the progressive decline in PBO levels in Olyset^®^ Plus and chlorfenapyr in Interceptor^®^ G2 highlights the importance of monitoring insecticide content degradation and its potential impact on long-term efficacy. The observed reduction in personal protection with Interceptor^®^ G2 over time further emphasizes the value of assessing multiple entomological outcomes, including both mortality and blood-feeding inhibition, when evaluating long-term ITN performance. These findings reinforce the necessity for extended longitudinal durability assessments conducted across diverse ecological and epidemiological settings in Africa to generate context-specific data that can inform ITN product selection, refine replacement timing, and support data-driven policy and procurement decisions in malaria vector control.

## Data Availability

All relevant data are within the article and its Supporting Information files.

## List of abbreviations

PBO: Piperonyl butoxide
ITN: Insecticide-treated nets
LLIN: Long-lasting insecticidal nets
WHO: World Health Organization
PQ: Prequalification team
AI: Active ingredient
cRCT: Cluster randomised controlled trial
SSA: Sub Saharan Africa
CREC: Centre de Recherche Entomologique de Cotonou
AIRID: African Institute for Research in Infectious Diseases
PAMVERC: Pan African Malaria Vector Research Consortium

## Declarations

### Ethical approval and consent to participate

Ethical approval was obtained from obtained from the Comité Nacional de Bioética para a Saúde em Moçambique (Ref:/424/CNBS/20) of Mozambique for the withdrawal field-aged nets was obtained from and the ethics review board of the Ministry of Health (CNERS) for the hut trial. Informed written consent was obtained from all householders and human volunteer sleepers before their participation study. A stand-by nurse was available throughout the trial to assess any volunteers presenting with febrile symptoms or an adverse reaction to the test items. The methods described in this paper followed relevant guidelines and regulations. Approval for using guinea pigs for tunnel tests was granted by LSHTM Animal Welfare Ethics Review Board (AWERB) (2020-01B).

### Consent for publication

Not applicable

### Authors’ contributions

M.R., J.W., H.K. and C.N. conceived the study. Field sampling of aged nets from households in Mozambique was led by APA and BC with support from H.K.. J.F. and B.N. performed the laboratory bioassays and hut trial. D.T. ensured the availability of test mosquitoes and performed the susceptibility bioassays. C.N. designed and supervised the laboratory bioassays and hut trial. M.B. and O.P. performed the chemical analysis. GGP and CF provided administrative and programmatic support. C.N. and J.F. analysed the data, prepared the figures and tables, and drafted the manuscript. All authors critically reviewed and approved the final version of the manuscript.

## Acknowledgements

We thank Binete Savaio from PATH Mozambique and the communities of the Gurue, Changara, Mandimba, and Guro districts of Mozambique and the rice farmers of Cove, Benin for their support and participation in the study. We appreciate the entire staff team of the CREC/PAMVERC laboratory team for their support.

